# *SMARCA4* Mutations in *KRAS*-mutant Lung Adenocarcinoma: A Multi-cohort Analysis

**DOI:** 10.1101/2020.06.27.20135020

**Authors:** Liang Liu, Tamjeed Ahmed, W. Jeffrey Petty, Stefan Grant, Jimmy Ruiz, Thomas W. Lycan, Umit Topaloglu, Ping-Chieh Chou, Lance D. Miller, Gregory A. Hawkins, Martha A. Alexander-Miller, Stacey S. O’Neill, Bayard L. Powell, Ralph B. D’Agostino, Reginald F. Munden, Boris Pasche, Wei Zhang

## Abstract

**Background:** *KRAS* is a key oncogenic driver in lung adenocarcinoma (LUAD). Chromatin-remodeling gene *SMARCA4* was co-mutated with *KRAS* in LUAD; however, the impact of *SMARCA4* mutations on clinical outcome has not been adequately established. This study sought to shed light on the clinical significance of *SMARCA4* mutations in LUAD.

**Methods:** The association of *SMARCA4* mutations with survival outcomes was interrogated in 4 independent cohorts totaling 564 patients: *KRAS*-mutant patients with LUAD who received non-immunotherapy treatment from 1) The Cancer Genome Atlas (TCGA) and 2) the MSK-IMPACT Clinical Sequencing (MSK-CT) cohorts; and *KRAS*-mutant patients with LUAD who received immune checkpoint inhibitor-based immunotherapy treatment from 3) the MSK-IMPACT (MSK-IO) and 4) the Wake Forest Baptist Comprehensive Cancer Center (WFBCCC) immunotherapy cohorts.

**Results:** Of the patients receiving non-immunotherapy treatment, in the TCGA cohort (n=155), *KRAS*-mutant patients harboring *SMARCA4* mutations (KS) showed poorer clinical outcome (*P*=6e-04 for disease-free survival (DFS) and .031 for overall survival (OS), respectively), compared to *KRAS*-*TP53* co-mutant (KP) and *KRAS*-only mutant (K) patients; in the MSK-CT cohort (n=314), KS patients also exhibited shorter OS than KP (*P*=.03) or K (*P*=.022) patients. Of patients receiving immunotherapy, KS patients consistently exhibited the shortest progression-free survival (PFS; *P*=.0091) in the MSK-IO (n=77), and the shortest PFS (*P*=.0026) and OS (*P*=0.0014) in the WFBCCC (n=18) cohorts, respectively.

**Conclusions:** mutations of *SMARCA4* represent a genetic factor that lead to adverse clinical outcome in lung adenocarcinoma treated by either non-immunotherapy or immunotherapy.

## BACKGROUNDS

Lung cancer is the leading cause of cancer-related death worldwide, with 5-year survival rates of ∼18%. Non-small cell lung cancer (NSCLC) comprises 85% of all lung cancer cases, mainly including adenocarcinoma (LUAD), squamous cell carcinoma (LUSC), and large cell carcinoma. Great strides have been made in recent years with the development of immune checkpoint inhibitor treatment targeting PD-1/PD-L1 mediated immunosuppression, which have shown efficacy in up to 30% of NSCLC patients [1-6]. Expression of PD-1/PD-L1 was reported to be associated with enhanced benefits from immunotherapy, but debates exist because of discordant results across different studies [1, 2, 4-11]. Currently, a higher tumor mutation burden (TMB) is undergoing evaluation as a predictive biomarker in many tumor types [7, 12-14].

The mutations in *KRAS* are a common oncogenic driver in ∼20% NSCLC [15, 16]. The goal of developing specific therapeutic strategies for the *KRAS*-mutant patients has thus far proven elusive. For example, *KRAS* mutations are associated with shortest survivals in NSCLC patients treated with carboplatin plus paclitaxel as well as single anti-EGFR TKI agent [17]. Recently, it was shown that *STK11*/*LKB1* or *TP53* co-mutations can stratify *KRAS*-mutant LUAD patient into different subgroups with distinct biology, therapeutic vulnerabilities and immune profiles [18], and immunotherapy response [19].

The SWItch/Sucrose NonFermentable (SWI/SNF) complex is a major chromatin remodeling complex that controls DNA accessibility to transcriptional factors and regulates transcriptional programming [20]. Genomic alterations in the components of the SWI/SNF chromatin remodeling complex have been identified in multiple types of cancers [21]. A recent study reported that mutations in the chromatin remodeling gene *PBRM1* were associated with response to immunotherapy through IFN-γ signaling pathway, a key effector for antitumor T cell function, in clear cell renal cell carcinoma [22, 23]. Mutations in the *PBRM1* in NSCLC is rare; however, mutations in the *SMARCA4* gene occur frequently in NSCLC [16, 24] and tended to co-occur with *KRAS* mutations [16]. One recent study showed that *SMARCA4* acted as a tumor suppressor by cooperating with p53 loss and Kras activation, and *SMARCA4*-mutant tumors were sensitive to inhibition of oxidative phosphorylation [25]. Another study showed that the reduced expression of *SMARCA4* contributes to poor outcomes in lung cancer [26]. However, the prognostic values of *SMARCA4* mutations in *KRAS*-mutant LUAD patients who received either non-immunotherapy or immunotherapy treatment have not been well defined.

In this study, we evaluated the prognostic value of *SMARCA4* mutations in *KRAS*-mutant LUAD within four independent cohorts consisting of patients received non-immunotherapy or immunotherapy treatment.

## METHODS

For the Cancer Genome Atlas (TCGA) cohort, matched somatic mutation, gene expression and clinical data of 560 patients with LUAD were retrieved. We obtained the clinical and somatic mutation data of 62 principal tumor types for MSK-IMPACT Clinical Sequencing Cohort and extracted the data of LUAD patients [27]. We excluded patients who received immunotherapy treatment indicated in their later publication [14] (as the MSK-IO cohort including 186 patients) to establish an MSK-CT cohort of 1033 patients received non-immunotherapy treatment.

We extracted the 127 LUAD patients who were treated with immunotherapy between March 1, 2015 and November 30, 2017 at the Wake Forest Baptist Comprehensive Cancer Center (WFBCCC) immune-oncology program. Efficacy was assessed by the treating physician and categorized according to RECIST guidelines [28] and defined as durable clinical benefit (DCB; complete response [CR]/partial response [PR] or stable disease [SD] that lasted > 6 months) or no durable benefit (NDB, PD or SD that lasted ≤ 6 months). Progression-free survival (PFS) was defined as the time from the date of initial immunotherapy administration to the date of progression or death, and overall survival (OS) was to the date of death or last follow-up, respectively. If the patient was alive at the date of last contact, his/her data were censored at that time point. Genomic profiles were available for 39 patients who were enrolled into the Wake Forest Precision Oncology Initiative (ClinicalTrials.gov Identifier: NCT02566421).

Only patients harboring *KRAS* mutations and with survival data were included in the study, resulting in 155 (27.7% of 560) and 314 (30.4% of 1033) patients received non-immunotherapy treatment in the TCGA and MSK-CT cohorts, and 77 (41.4% of 186) and 18 (46.2% of 39) patients received immunotherapy treatment in the MSK-IO and the WFBCCC cohorts.

### Statistical Analysis

Tests used to analyze clinical and genomic data included the Mann-Whitney U test (two-group comparisons), χ^2^ test (three-group comparisons), Fisher’s exact test (proportion comparisons). Survival curves were estimated using Kaplan-Meier methodology and compared between two groups using the log-rank test and Cox proportional hazards regression analysis. Hazard ratios (HRs) and 95% CIs were generated by Cox proportional hazards models where *P*<.05 and these statistics were estimable (i.e., when at least one event occurred in both groups being compared). All analyses were performed using R software, version 3.2.1.

## RESULTS

### *SMARCA4* Mutations are Associated with Shorter Survival of Patients Who Received Non-immunotherapy Treatment

*KRAS* is one of the most frequently mutated genes in LUAD, which occur in 155 (30%) patients in the TCGA cohort. These patients were reprehensive of the overall LUAD cohort with median patient age of 67 years (range 33-87) and high percentage of current/former smokers (94.8%). 5.8% (9) of the *KRAS*-mutant patients harbored *SMARCA4* mutations in the TCGA cohort and were classified as KS; 33.5% (52) patients harbored *TP53* mutations and were classified as the KP subgroup; and 60.6% (94) patients did not carry *SMARCA4* or *TP53* mutations and were classified as K (**Supplementary Figure 1**). The *SMARCA4* mutations were not associated with any risk factors such as age at diagnosis, tumor stage, race/ethnicity or smoking history (**Supplementary Table 1**).

Disease-free survival (DFS) differed between the three groups (*P*=6e-4), with significantly shorter DFS for patients in the KS subgroup compared to either KP (HR 4.47, 95% CI 1.52-13.22, *P*=.003) or K (HR 2.43 95% CI 1.46-4.05, *P*=1.2e-4) patients in pair-wise comparisons (**Figure 1A**). In contrast, KP and K patient had similar DFS (*P*=.64). We also compared the survivals between KS (*SMARCA4*-mutant) and KP+K (*SMARCA4*-wildtype) patients, and found that KS patients exhibited significantly shorter DFS (HR 5.34 95% CI 2.05-14.14, *P*=1.3e-4) (**Figure 1B**).

**Figure 1:**
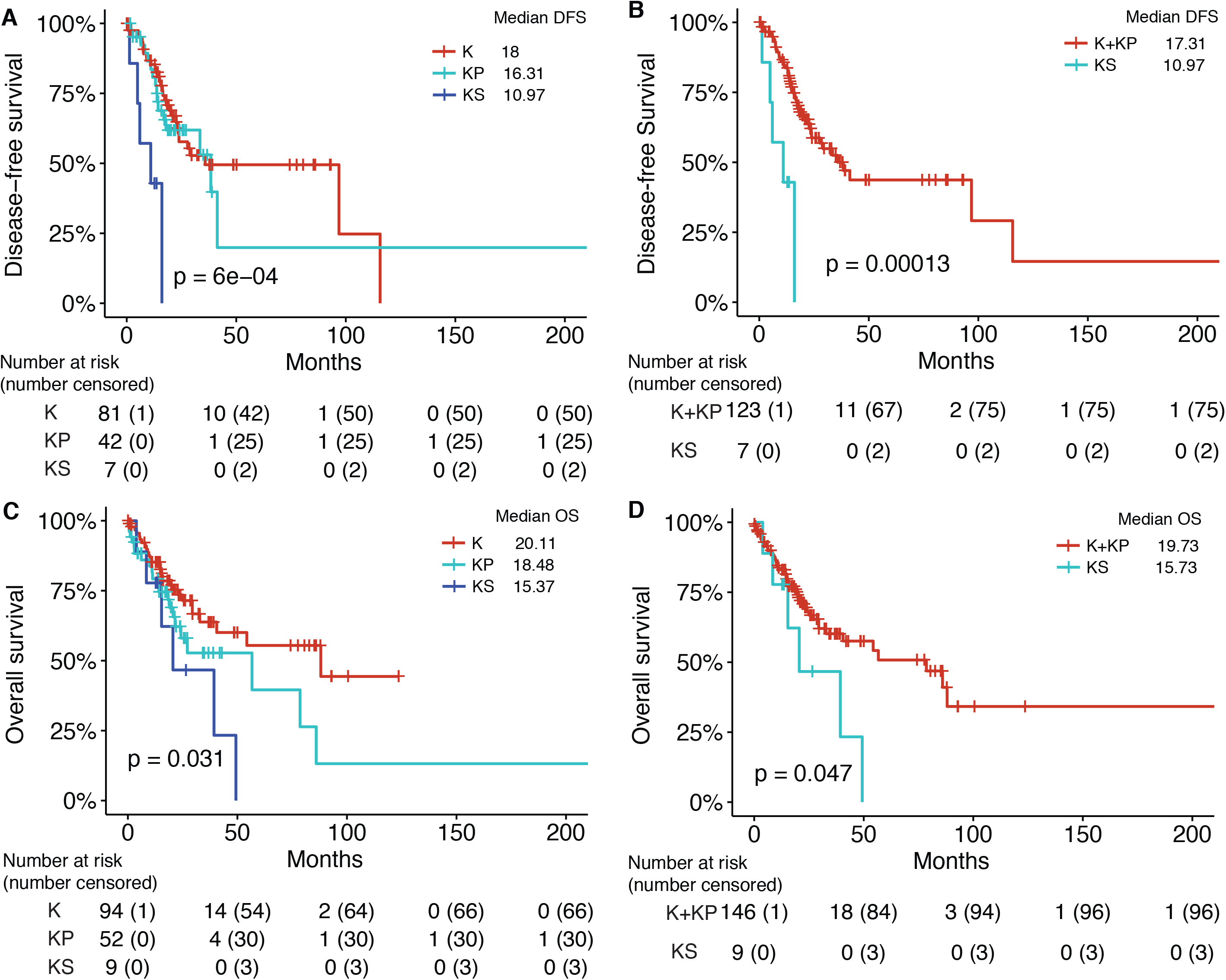
*SMARCA4* mutations are associated with shorter disease-free survival (DFS) and overall survival (OS) of *KRAS*-mutant LUAD patients treated with non-immunotherapy treatment from the TCGA cohort. Kaplan-Meier survival analysis of survival in (AC) the KS, KP and K subgroups and (BD) in the two-group comparison between *SMRACA4*-mutant and wildtype *KRAS*-mutant patients.

Overall survival (OS) also varied significantly between the three groups (*P*=.031). The KS patients exhibited shorter DFS than the K subgroup (HR 1.63, 95% CI 1.05-2.55, *P*=.024). Although the difference of OS between KS and KP was not significant (*P*=.21), the median OS in KS was 15.37 months compared to 18.48 months in KP (**Figure 1C**). In addition, the two-group comparison showed significantly shorter OS in KP (*SMARCA4*-mutant) compared to K+KP (*SMARCA4*-wildtype) patients (HR 2.32, 95% CI 1.01-5.44, *P*=.047) (**Figure 1D**).

We validated these observations in an independent MSK-CT cohort [27], consisting of 314 *KRAS*-mutant patients. High percentage of current/former smokers (78.0%) were also observed. Across the entire cohort, 10.8% (34) patients were classified as KS, 34.1% (107) were KP and 55.1% (173) were K (**Supplementary Figure 1 and Table 2**). Significantly shorter OS were observed for patients with KS compared to K (HR 1.39, 95% CI 1.04-1.85, *P*=.022) or KP (HR 1.94, 95% CI 1.06-3.57, *P*=.03) (**Figure 2A**), and K and KP have similar OS (*P*=.99). In the two-group comparison, OS was significantly shorter in KS (*SMARCA4*-mutant) compared to K+KP (*SMARCA4*-wildtype) patients (HR 1.95, 95% CI 1.13-3.38, *P*=.015) (**Figure 2B**).

**Figure 2:**
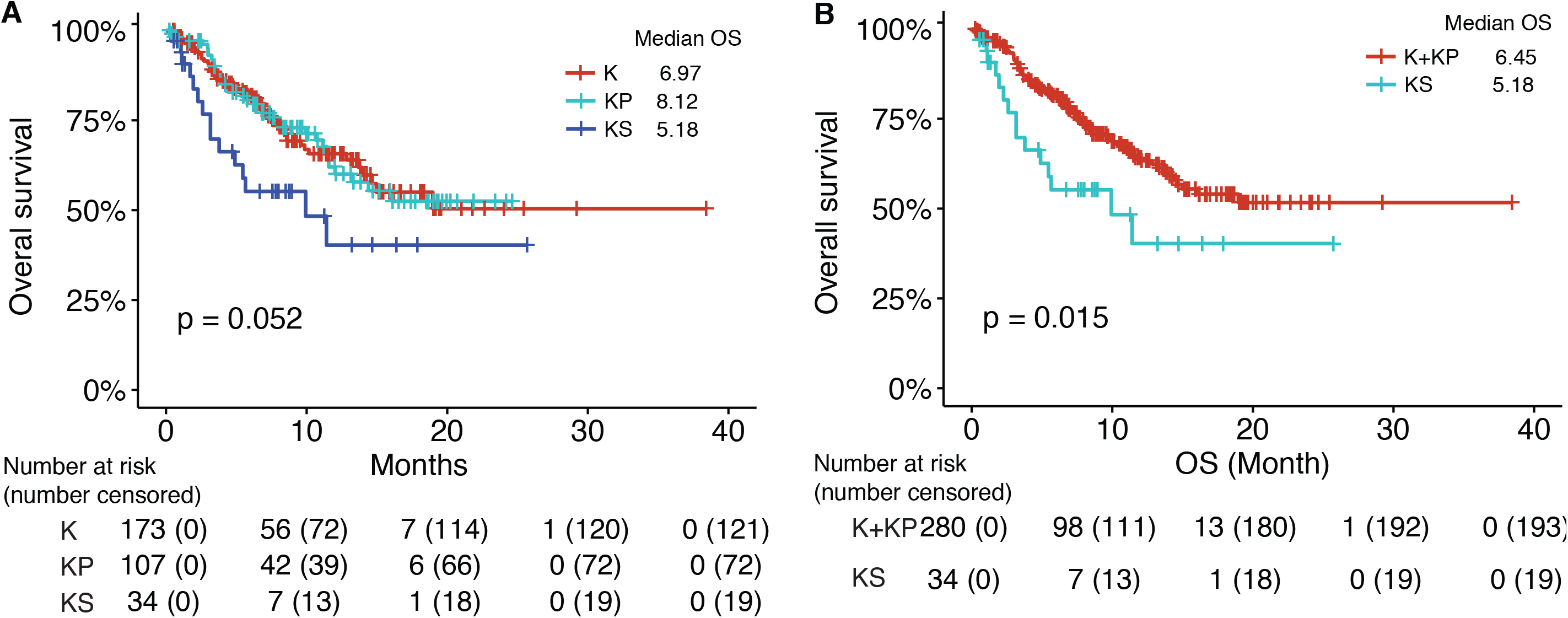
*SMARCA4* mutations are associated with shorter overall survival (OS) of *KRAS*-mutant LUAD patients treated with non-immunotherapy treatment from the MSK-CT cohort. Kaplan-Meier survival analysis of OS (A) in the KS, KP and K subgroups and (B) in the two-group comparison between *SMRACA4*-mutant and wildtype *KRAS*-mutant patients.

### *SMARCA4* Mutations are Associated with Shorter Survival of Patients Who Received Immunotherapy Treatment

We then examined whether *SMARCA4* mutations impacted *KRAS*-mutant patient response to immunotherapy. 77 LUAD patients harboring *KRAS* mutations were extracted from the MSK-IO cohort [14]. The median age of patients was 68 (range 37-86) and the majority (93.5%) was ever smokers. Based on *SMARCA4* and *TP53* mutation status, 11.7% (9) tumors were classified as KS, 32.5% (25) were KR, and 55.8% (43) were K. Demographic and clinical characteristics were generally well balanced between the co-mutation defined groups. The clinical benefit rates to immunotherapy in KS, KP and K groups was not significantly different (*P*=.42), probably due to the small sample size; however, smaller proportion of KS patients (2/9=22.2%) achieved durable clinical benefit (DCB) than KP (10/23=43.5%) or K (13/43=30.2%) patients. (**Supplementary Figure 1 and Table 3**).

Significantly different PFS was observed between the three groups (*P*=.0091). The KS patients exhibited the shorter PFS compared to KP (HR 2.82, 95% CI 1.17-6.81, *P*=.016) tumors in pair-wise comparisons. Although the difference of PFS between KS and K was not significant (*P*=.18), the median OS in KS was 1.73 months compared to 2.77 months in KP. Interestingly, KP patients exhibited longer survival than K patients (HR 0.48, 95% CI 0.26-0.86, *P*=.012) (**Figure 3A**). We merged the KP and K patients to test the difference between *SMARCA4*-mutant and wildtype patients. *SMARCA4*-mutant (KS) patients exhibit significantly shorter PFS than wildtype (K+KP) patients (HR 2.15, 95% CI 1.46-4.35, *P*=.048, median PFS 1.73 vs. 4.22 months) (**Figure 3B**).

**Figure 3:**
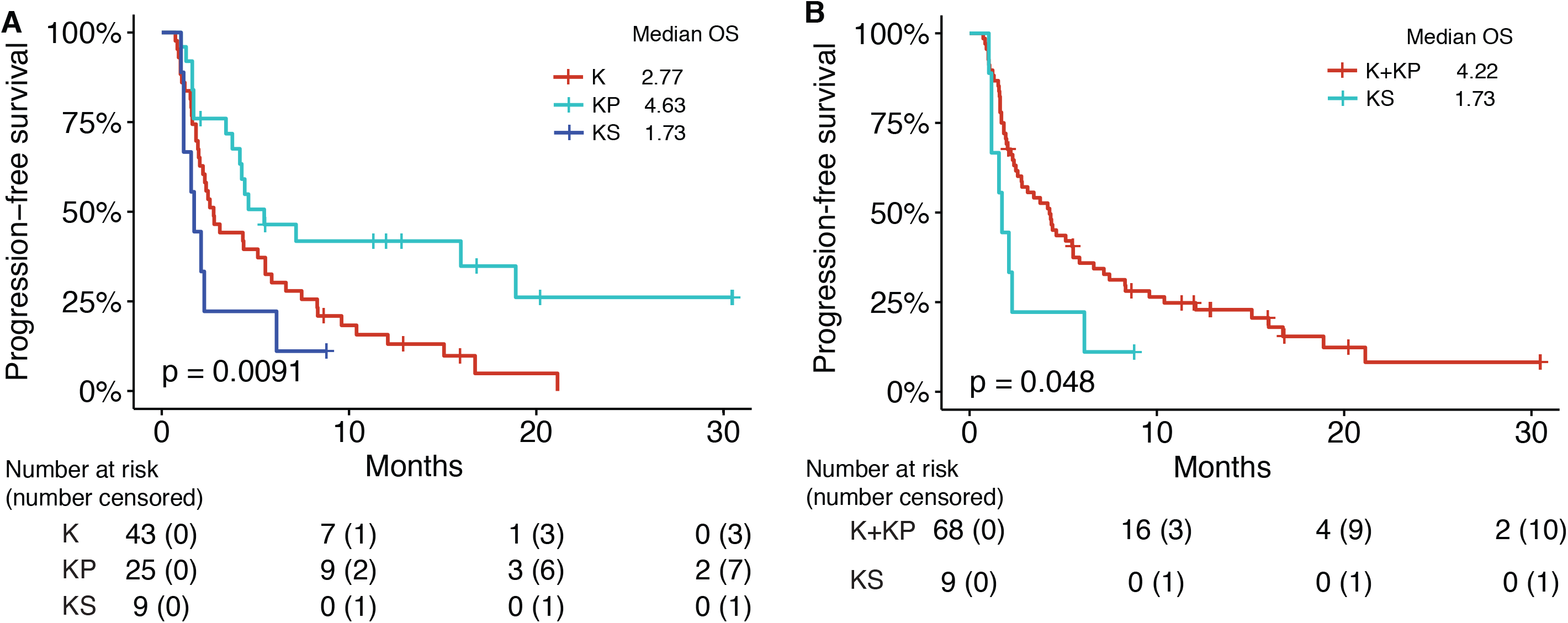
*SMARCA4* mutations are associated with shorter progression-free survival (PFS) of *KRAS*-mutant LUAD patients treated with immunotherapy treatment from the MSK-IO cohort. Kaplan-Meier survival analysis of PFS (A) in the KS, KP and K subgroups and (B) in the two-group comparison between *SMRACA4*-mutant and wildtype *KRAS*-mutant patients.

We also validated the prognostic values of *SMARCA4* mutations in *KRAS*-mutant LUAD patients upon immunotherapy using 18 patient samples from the WFBCCC. Patients were classified into KS (11.1%), KR (44.4%) and K (44.4%) subgroups (**Supplementary Figure 1 and Table 4**). In this small cohort, the clinical benefit rates to checkpoint inhibitor-based immunotherapy in KS, KP and K groups were significantly different (*P*=.03). KS patients were resistant to treatment, while KP patients were mostly sensitive.

The three groups of *KRAS*-mutant LUAD patients exhibited significantly different OS (*P*=.042) and PFS (*P*=.0014). The KS patients exhibited the shortest OS and PFS compared to either KR (HR 2.46, 95% CI 1.05-6.61, *P*=.0019 and *P*=.0019 with HR and 95% CI evaluable) and K (HR 2.46, 95% CI 1.01-6.61, *P*=.042 and HR 3.06, 95% CI 1.03-10.28, *P*=.029) patients in pair-wise comparisons (**Figures 4A and 4C**). Further significantly deceased OS and PFS was observed in KS (*SMARCA4*-mutant) patients compared to K+KP (wildtype) ones (HR 11.98, 95% CI 1.66-26.6, *P*=.0018 and HR 18.7, 95% CI 1.65-21.6, *P*=.0011) (**Figures 4B and 4D**), consistent with the observations in the MSK-IO cohort. Altogether, these data indicated that *SMARCA4* abrogation likely determines immunotherapy resistance in *KRAS*-mutant LUAD.

**Figure 4:**
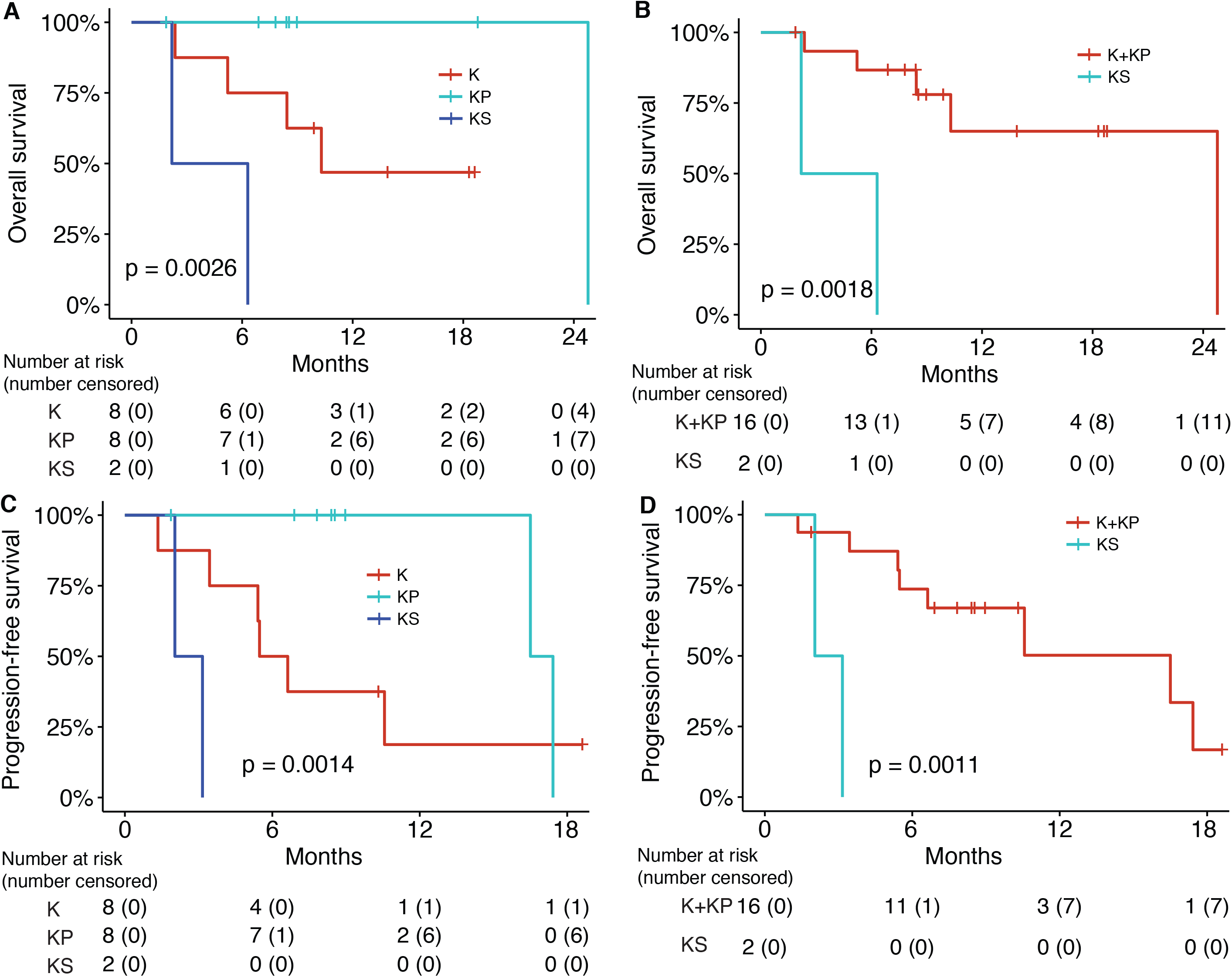
*SMARCA4* mutations are associated with shorter progression-free survival (PFS) and overall survival (OS) of *KRAS*-mutant LUAD patients treated with immunotherapy treatment from the WFBCCC cohort. Kaplan-Meier survival analysis of survival (AC) in the KS, KP and K subgroups and (BD) in the two-group comparison between *SMRACA4*-mutant and wildtype *KRAS*-mutant patients.

### *SMARCA4* Mutations are Significantly Enriched among Tumors with Immunosuppressive Tumor Microenvironment

We interrogated the composition of immune cells in the tumor microenvironment of patients from the TCGA cohort which has RNA-seq data available. Using CIBERSOFT [29] to quantify the proportion of each individual immune cell type, we found that KS patients had significantly lower estimated proportions of CD8 and activated CD4 memory T cells than either K (*P*=.015 and .035) or KS (*P*=.043 and .023), while no differences between KP and K patients (*P*=.66 and .35), indicating an immunosuppressive tumor microenvironment in the KS patients (**Figure 5 and Supplementary Figure 2**).

**Figure 5:**
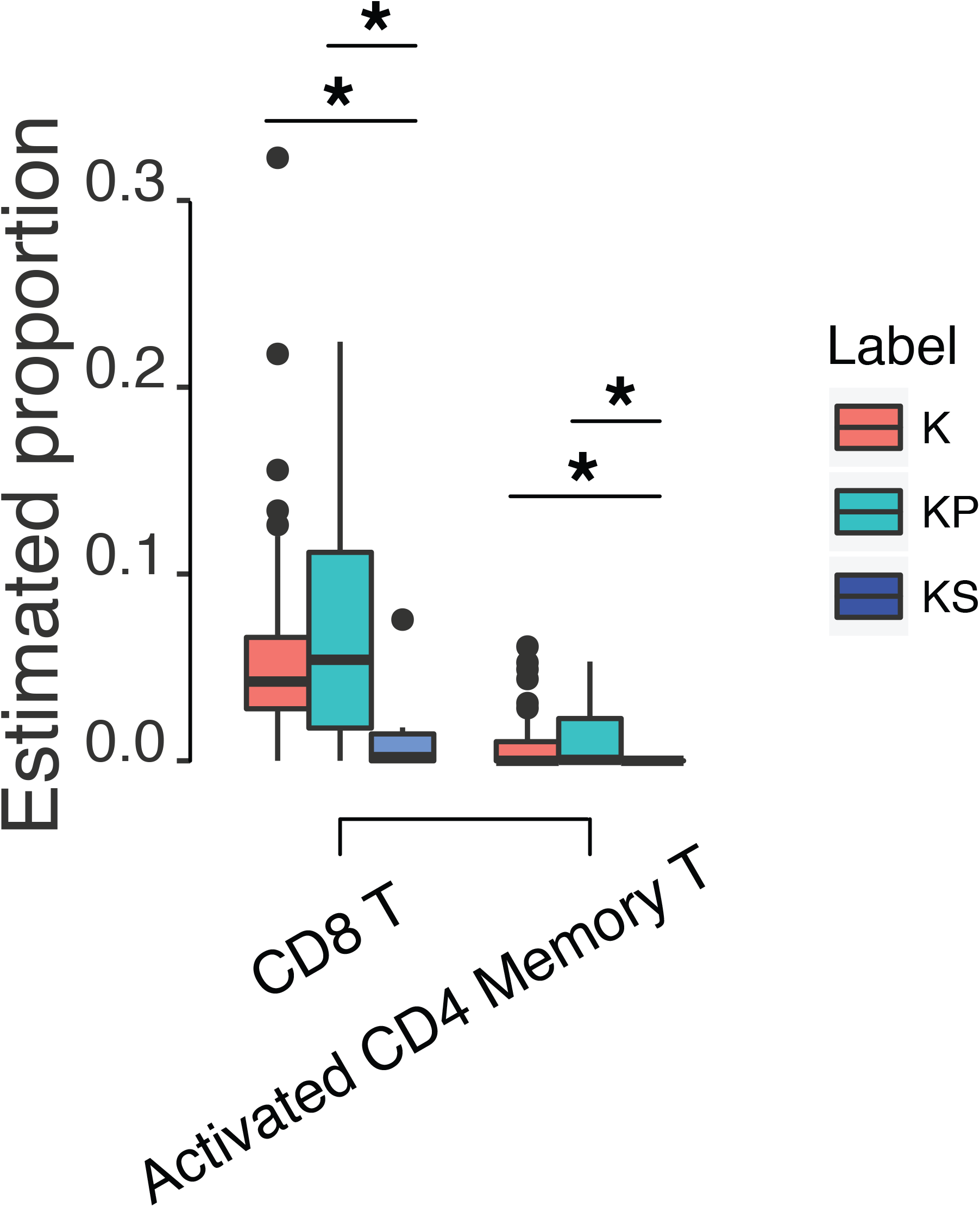
Tumor microenvironment varied among three groups of patients. KS patients contained the lowest proportions of CD8 and activated CD4 memory T cells than either K or KP patients. The plot for all 22 types of immune cells was shown in **Supplementary Figure 2**. \**P*<.05; Mann-Whitney U test.

## DISCUSSION

Alterations in chromatin remodeling complex, SWI/SNF, including *SMARCA4*, have been found in NSCLC [16, 24, 30, 31]. In this study, we interrogated the clinical significance of *SMARCA4* mutations in *KRAS*-mutant LUAD in the TCGA and the MSK-CT cohorts in the absence of immunotherapy and the MSK-IO and the WFBCCC cohorts who received immunotherapy. Our analysis indicates that genomic alterations in the chromatin remodeling gene, *SMARCA4*, as a negative prognostic factor to *KRAS*-mutant LUAD patients no matter received non-immunotherapy or immunotherapy treatment. The mutations may induce an immunosuppressive tumor environment by modulating the immune cell components. Although the completed determinants of response to treatment is not yet completed defined, our study suggests that non-immunotherapy and immune checkpoint inhibitor-based immunotherapy treatment may not benefit this subset of patients.

More frequent *KRAS* mutations were observed in ever smokers than that occurred in never smokers [24, 32-34], and associated with a significant increase of TMB [35]. Previous studies indicated that a subset of *KRAS*-mutant NSCLC patients may have a better response to immunotherapy treatment [2, 35, 36]. We determined that KP patients exhibited better survival than KS and K patients when receiving immune checkpoint inhibitor-based immunotherapy, which is consistent with previous report [35]. The underlying mechanism may be that KP patients contained the largest proportion of CD8 and activated CD4 memory T cells, supporting by previous report that *TP53* and *KRAS* mutations had remarkable effects on increasing *PD-L1* expression, facilitating T-cell infiltration and augmenting tumor immunogenicity [35].

*SMARCA4* inactivation was shown to promotes NSCLC aggressiveness by altering chromatin organization [30], and the reduced expression of *SMARCA4* contributes to poor outcomes in lung cancer [26]. Here we showed that *SMARCA4*-*KRAS* co-mutant patients (KS) exhibited poorer survival of patients who received either non-immunotherapy or immunotherapy treatment. On the other hand, quantitative IHC for BRG1 can capture *SMARCA4*-deficient tumor [37, 38] which is associated with *SMARCA4* mutations (**Supplementary Figure 3**). Therefore, evaluation of BRG1 expression by IHC may further enhance the predictive utility for non-immunotherapy or immunotherapy treatment to NSCLC.

*SMARCA4* mutation is a unique biomarker for the stratification of *KRAS*-mutant patients with LUAD. SMARCA4 mutations are not associated with other factors such as *STK11/LKB1* mutations, which can stratify *KRAS*-mutant LUAD into different subgroups with distinct biology, therapeutic vulnerabilities and immune profiles [18] and immunotherapy response [19], because mutations in *SMARCA4* and *STK11/LKB1* did not co-occur in *KRAS*-mutant patients receiving immunotherapy treatment in the MSK-IO (*P*=0.065) or WFBCCC (*P*=0.41) cohorts. In addition, *STK11* mutations did not serve as a prognostic marker for patients who received non-immunotherapy treatment [19, 39-41].

For these patients harboring both *KRAS* and *SMARCA4* mutations, an alternative treatment strategy is required. A clinical study showed that cisplatin-based chemotherapy benefited NSCLC patients with low *SMARCA4* expression [26]. Another report indicated the activity of *AURKA*, which encodes a cell-cycle regulated kinase, was essential in NSCLC cells lacking *SMARCA4*, and the inhibition/depletion of *AURKA* enabled apoptosis and cell death *in vitro* and in xenograft mouse models [42]. Moreover, a recent study indicated that *SMARCA4*-deficient lung cells and xenograft tumors displayed marked sensitivity to inhibition of oxidative phosphorylation [25]. All observations suggested encouraging treatment strategies but need further testing in clinics.

## CONCLUSIONS

We provide evidence that *SMARCA4* mutations are associated with poor clinical survival outcomes of *KRAS*-mutant LUAD patients. If confirmed in additional cohorts, it is likely that future prediction models will need to include *SMARCA4* mutations.

## Data Availability

All relevant data and materials within this work are made available in this manuscript or previously published.

## ABBREVIATIONS

LUAD: Lung adenocarcinoma
KS: KRAS-SMARCA4 co-mutant
KP: KRAS-TP53 co-mutant
K: KRAS-only mutant
LUSC: Lung squamous carcinoma
NSCLC: Non-small cell lung cancer
TCGA: The Cancer Genome Atlas (TCGA)
MSK-CT: the MSK-IMPACT Clinical Sequencing cohort
MSK-IO: MSK-IMPACT cohort
WFBCCC: the Wake Forest Baptist Comprehensive Cancer Center
DCB: durable clinical benefit

## Acknowledgement

W.Z. is supported by the Hanes and Willis Professorship in Cancer and a Fellowship from the National Foundation for Cancer Research. B.P. is supported by the Charles L. Spurr Professorship Fund. We thank Dr. Matthew S. Hellman at the MSKCC for helpful discussion of the results especially in regard to the MSK-IO cohort. We also acknowledge the editorial assistance of Karen Klein, MA, in the Wake Forest Clinical and Translational Science Institute, and Dr. Mac Robinson at the WFBCCC.

## Authors’ contributions

All authors have participated in drafting, reading, and approving the final manuscript.

## Consent for publication

Not applicable.

## Ethics approval and consent to participate

Approval for the FoundationOne test were obtained from the ethics committee.

## SUPPLEMENTAL CONTENT

**Supplementary Table 1:** *KRAS*-mutant patient Characteristics in the TCGA cohort treated with non-immunotherapy.

**Supplementary Table 2:** *KRAS*-mutant patient Characteristics in the MSKCC-CT cohort treated with non-immunotherapy.

**Supplementary Table 3:** *KRAS*-mutant patient Characteristics in the MSK-IO cohort treated with immunotherapy.

**Supplementary Table 4:** *KRAS*-mutant patient Characteristics in the WFBCC cohort treated with immunotherapy.

**Supplementary Figure 1:** Global somatic mutation landscape of *KRAS, TP53* and *SMARCA4* genes in the TCGA, MSK-CT, MSK-IO and WFBCCC cohorts.

**Supplementary Figure 2:** The comparisons of estimated proportions of immune cell subsets, as calculated by CIBERSORT among K, KP and KS patients. Across all cell types, the proportions of CD8 T cells and activated CD4 memory T cells differ significantly in the comparisons of K vs. KS and KP vs. KS (shown in Figure 5).

**Supplementary Figure 3:** *SMARCA4* mutations are associated with lower expression level of *SMARCA4*.

